# Dynamic Analysis of Social Distancing Ratio, Isolation Rate and Transmission Coefficient in COVID-19 Epidemic for Many Countries by SIQR Model

**DOI:** 10.1101/2020.08.04.20167882

**Authors:** Koichi Hashiguchi

## Abstract

Recently SIQR model was proposed by Odagaki as the modification of conventional SIR model by adding the term for isolation of infected persons, Q(Quarantined). The exponent *λ* of the exponential function expressing the number of newly tested positive persons was defined as an linear equation explicitly with three important parameters, transmission coefficient, social distancing ratio x and isolation rate q. In this study, applying this model to the number of positive persons in publicly available database, daily *λ* values are regression analyzed, and social distancing ratio and isolation rate are derived. Analyses for 7 countries including Japan, Taiwan, South Korea, and western countries are performed and determine the dynamic locus of q-x relation on the q-x plane during epidemic propagation. Finally, the remaining parameter, the transmission coefficient is shown to closely relate to the maximum λ, λ_max_, and λ_max_ (transmission coefficient) is characterized as a specific value for each country. Then, the magnitude of λ_max_ is combined with the value of λ_min_ to influence the total number of new cases until the convergence stage.

## Introduction

A new type of coronavirus, COVID-19, which took off in Wuhan, China, at the end of 2019 and quickly spread throughout the world, has not subsided even after six months, and is showing signs of further spreading.

The SIR model ^1)^ proposed about 100 years ago is known as a mathematical model for describing the epidemic process of infection. This model reproduced the plague epidemic of the early 20th century and is also believed to help understand the current COVID-19 epidemic.^2)^ In 2020, Odagaki proposed the SIQR model ^3)^ in which the recovered group, R, in the SIR model was divided into the quarantined, Q and the recovered, R. With additional group of uninfected, S (Susceptible), and community infected, I (Infected), the in and out of the number of people between each group were expressed in the differential equations with the overall condition of preservation of population. Three important parameters, transmission coefficient, isolation rate and social distancing ratio, which are essential for infection control, were explicitly introduced to bring more concreteness to the expression of actual infection status and it is expected to contribute to evaluating the infection control.

In order to evaluate the validity of the model and to clarify the status of infection in several countries, the number of daily isolated persons(new cases) in a publicly available database are analyzed based on the SIQR model, and the social distancing ratio and isolation rate are derived. Then the infection process in each country from the initial stage to the convergence stage are characterized by using these two parameters. Finally, the relationship between the number of new cases until the convergence of infection and the transmission coefficient, which is the third important parameter as specific value for each country in this model, are discussed.

## Analyzing Method

### Basic Equations in SIQR model and Matching Method in data-fitting

In the SIQR model by Odagaki, the daily variation in the number of persons in each group of S, I, Q, and R is represented by four differential equations, and λ, corresponding to the exponent after integration, was defined in Equation (1), with combining social distancing ratio x, isolation rate q and cure rate γ.

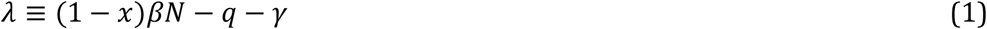

βN is the transmission coefficient β multiplied by population N, which is an index of the degree of transmission through contact between uninfected and infected individuals. Since x and q are both time-dependent parameters, exact integration of the above mentioned differential equations are difficult. Therefore, Odagaki divided the infection period into the four stages, first stage of the initial rise of the infection, the second stage of the rapid increase period, the third stage of the plateau with the maximum value, and the fourth stage of the convergence. Assuming that the number of daily quarantined persons (tested as positive and maybe isolated, hereafter new cases) ΔQ for each stage is represented by an exponential function with a different exponent, ΔQ and the number of infected persons in the community I are expressed by equations (2) and (3), respectively. Then Odagaki analyzed the social distancing ratio and the isolation rate separately for 4 stages.

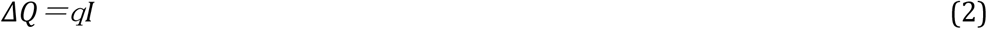

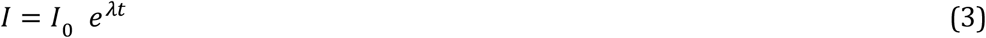

In this analysis, the same approximation method was adopted except Odagaki’s classification of the 4 stages which was replaced with a day-to-day basis.. The number of new cases (*ΔQ*) on a daily basis was expressed by an logarithmic approximate expression (4) with time unit of a day and constant term of *ΔQ*_0_, and the exponent thereof was expressed by eq. (1) to proceed with the analysis.

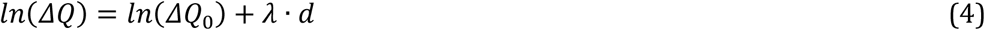

Daily *λ* were regression analyzed using eq. (4) with day range [*d_i_* − Δ*d*, *d_i_* + Δ*d*], where *d_i_* is the current day, and Δ*d* corresponds to day range. This procedure was carried out not in the 4 stages of Odagaki but in each day during the period, which leads to the dynamic capturing of variation of infection process.

In each of four groups of S, I, Q, and R, the degree of influence on infection spread may differ due to depopulation, regional differences such as age composition, and differences in countermeasures, even though, only the average behavior in each country was considered similarly with Odagaki’s method.

Both the social distancing ratio x and the isolation rate q in eq. (1) are parameters that take values of 0 to 1 based on their definitions. When both are 0, that is, without any measures to suppress infection, the right side of eq. (1) becomes βN−γ, which is the maximum value λ_max_ (= βN−γ), and the number of new cases increases at the fastest speed. This relationship is substituted into eq. (1) and transformed to obtain equation (5).

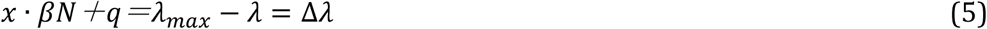

λ_max_ is considered constant throughout the period of infection. γ is the cure rate of community-infected individuals and is set at 0.04, following Makino ^3)^, and is not significantly influential during the following analysis. The units of λ, q and βN are same as (1/day), and x is dimensionless.

From equations (1) and (4), the daily new cases ΔQ is expressed as an exponential function that takes varying daily λ as an exponent. With obtaining daily λ, the right side ∆λ of eq. (5) is determined, using λ_max_, the maximum value of λ during the entire period. By sharing the value between x and q on the left side, it is possible to determine the social distancing ratio x and the isolation rate q. Figure 1 shows the variation of daily λ value for the case of Japan as an example. The initial stage of infection starts from a small positive value of λ, and after fluctuation, λ rapidly increases and reaches the maximum value λ_max_, then after repeating a slight increase/decrease, it passes through 0 and converges to a negative value. The increase in λ after May was ignored in this analysis. In this figure, ∆λ at each day is equal to the left side of eq. (5). Equation (5) is an indefinite equation consisting of two independent variables of x and q (both in the range of 0 to 1), which are not related to each other, so that some kind of ingenuity is required to solve it.

**Fig. 1.**
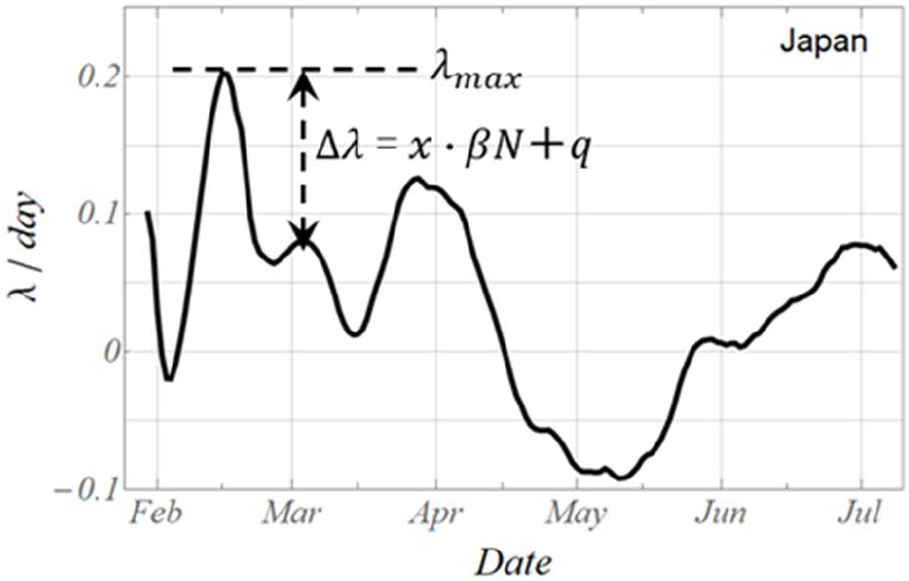
Schematic illustration of daily variation of exponent λ for Japan, explaining the way of fitting equation (5) in SIQR analysis

The terms frequently used throughout this paper are unified as follows. New infected and isolated persons as new cases, 1st and 2nd period by Odagaki’s paper by increasing stage, maximum number of new cases as peak, 4th period as converging stage, and the day of minimum number of new cases in the 4th period as the convergence day.

### Derivation of the social distancing ratio x and isolation rate q from the daily λ value (Solution of the indefinite equation)

The indefinite equation with unknown parameters q and x with substituting daily λ value to eq. (5) was solved in the following manner, under the rules that the initial conditions are given and that neither q nor x leads to an abrupt and large discontinuity.

At the start of infection, x was set to zero, and sequential calculations were performed using the following procedure.

On a first day; x_1_ = 0, and q_1_=λ_max_−λ_1_−x_1_ βN is calculated.
On the second day; set q_2_ = q_1_ and calculate x_2_ = (λ_max_−λ_2_−q_2_)/βN.
On the third day; x_3_=x_2_ and q_3_=λ_max_−λ_3_−x_3_ βN is calculated.

In this way, the sequential calculations were carried out by fixing one of q and x and moving the other, imitating a bipedal walk. Thus, on the q-x plane, the calculation starts on a q-axis (x=0) and follow the (q, x) coordinates in a stepwise manner afterwards. In the rare cases where x becomes negative due to the same value of q adopted from the previous day, x is set to 0 and q is calculated again.

### Data analysis of the number of new cases

For the analysis, the number of new cases in more than 200 countries in the “Our World in Data” database^4)^ (data coverage from 1^st^ of Jan. 2020 to 11^th^ of July) were used. There is a big difference in the daily data due to differences in the data aggregation method depending on the country, so a moving average was taken on a weekly basis. In addition, the number of new cases was converted to per million population for comparison by country.

In the calculation of daily λ value, the range of days for regression, 2Δ*d* +1 was varied from 7 days up to 31 days with varying Δ*d* as of 3, 9, 15. The period ofλ analysis was from the day when the cumulative number of new cases exceeds 10 persons, to the day when the number of new cases reaches the minimum after peak.

The following seven countries with different countermeasures were chosen for the analysis. Japan with the social distancing policy, Taiwan and South Korea that quickly settled due to swift response, Sweden for mass infection, Italy, Germany and the United States suffering from explosive mass outbreak.

## Dynamic Evolution of social distancing ratio and isolation rate

### Variation in the number of new cases by country and variation in daily λ

The daily variation of new cases and the trend curve of tdaily λ calculated with the regression range of 19 days for 7 countries are shown in Figure 2a and 2b, respectively. Also shown in Table 1 is the start day, the day for maximum of new cases (peak day) and the convergence day, and λ_max_ for these 7 countries. The maximum peak value (per million population) of the new cases is lowest in Taiwan, followed by Japan and South Korea, and the decrease in the number of new cases in western countries is small after the rapid increase to a higher level. The daily λ values exhibit rough variation along with the increasing number of new cases, with the result that λ_max_ is low in Taiwan and Japan.

**Fig. 2a,b.**
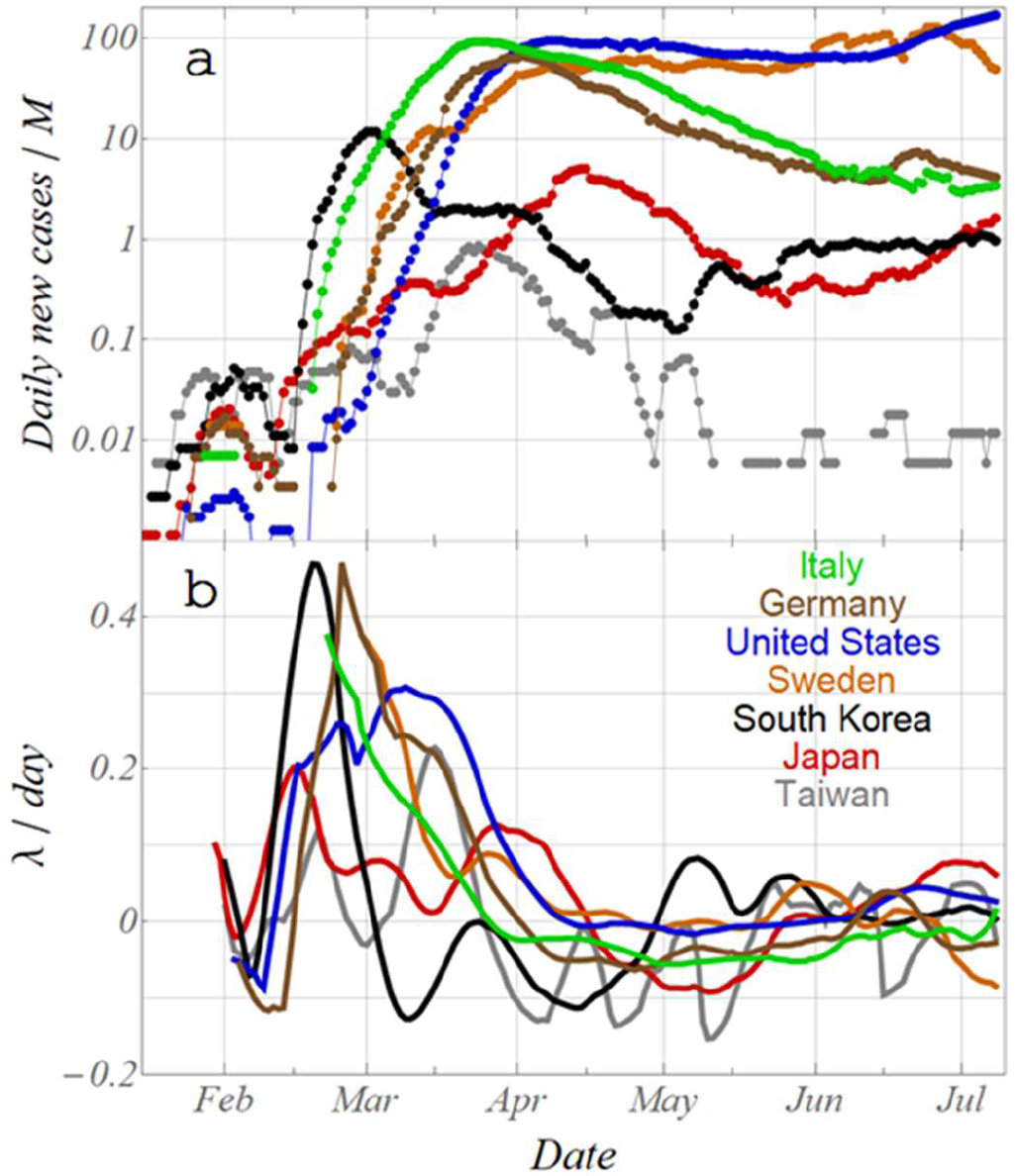
Variation of daily new cases and λ for 7 countries. Former are expressed as per million population for the comparison between countries.

**Table 1.**
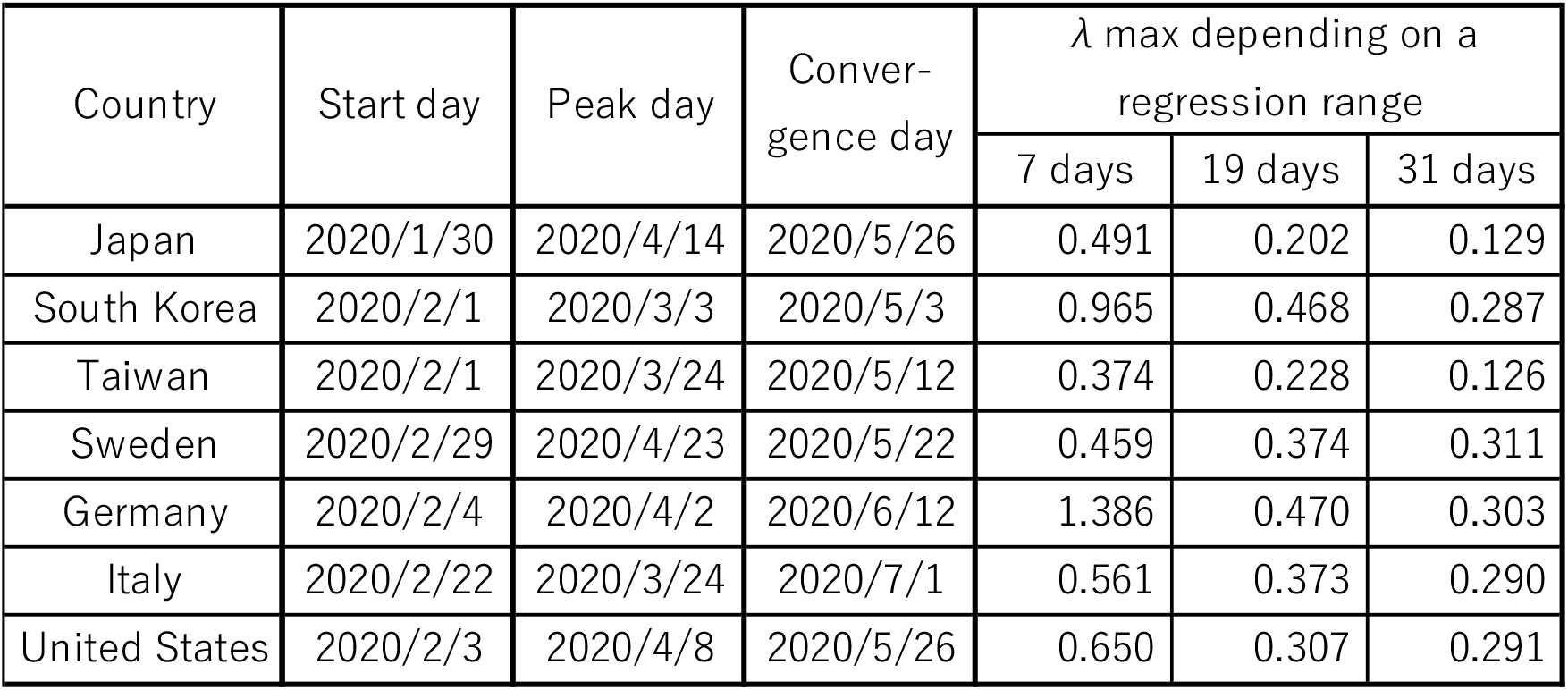
Remarked days during COVID-19 epidemic, and regression analyzed daily λ max dependance on a range of regression for 7 countries

Table 1 shows that the λ_max_ value varies greatly depending on the regression range *d_i_* ± Δ*d*, and as the range is expanded, the value becomes smaller due to the leveling-off effect. Hereafter, results for the range of λ calculation at 19 days with moderate leveling effect is discussed and the effect of the range will be discussed later.

### Characteristic variation of social distancing ratio x and isolation rate q for each country

Figures 3a and 3b show the evolution of the isolation rate q and the social distancing ratio x, respectively. Both parameters have stepwise varying curves due to the bipedal walk calculation. As a result of setting x to 0 on a first day as the initial condition, starting q ranged from 0.1 to 0.5, depending on the country-specific ∆λ as shown at the left end in Fig. 3a. After that, both q and x decrease once, and reach a minimum 0 at the day of λ_max_. Again, q and x increase with the increase in ∆λ, and after λ turns negative, q and x tend to converge to almost constant values respectively. Although the same tendency is shown in all countries, the absolute values of q and x depend on ∆λ, and the isolation rate of Germany and South Korea with higher λ_max_ are higher, while those of Japan with lower λ_max_ are lower.

**Fig. 3a,b.**
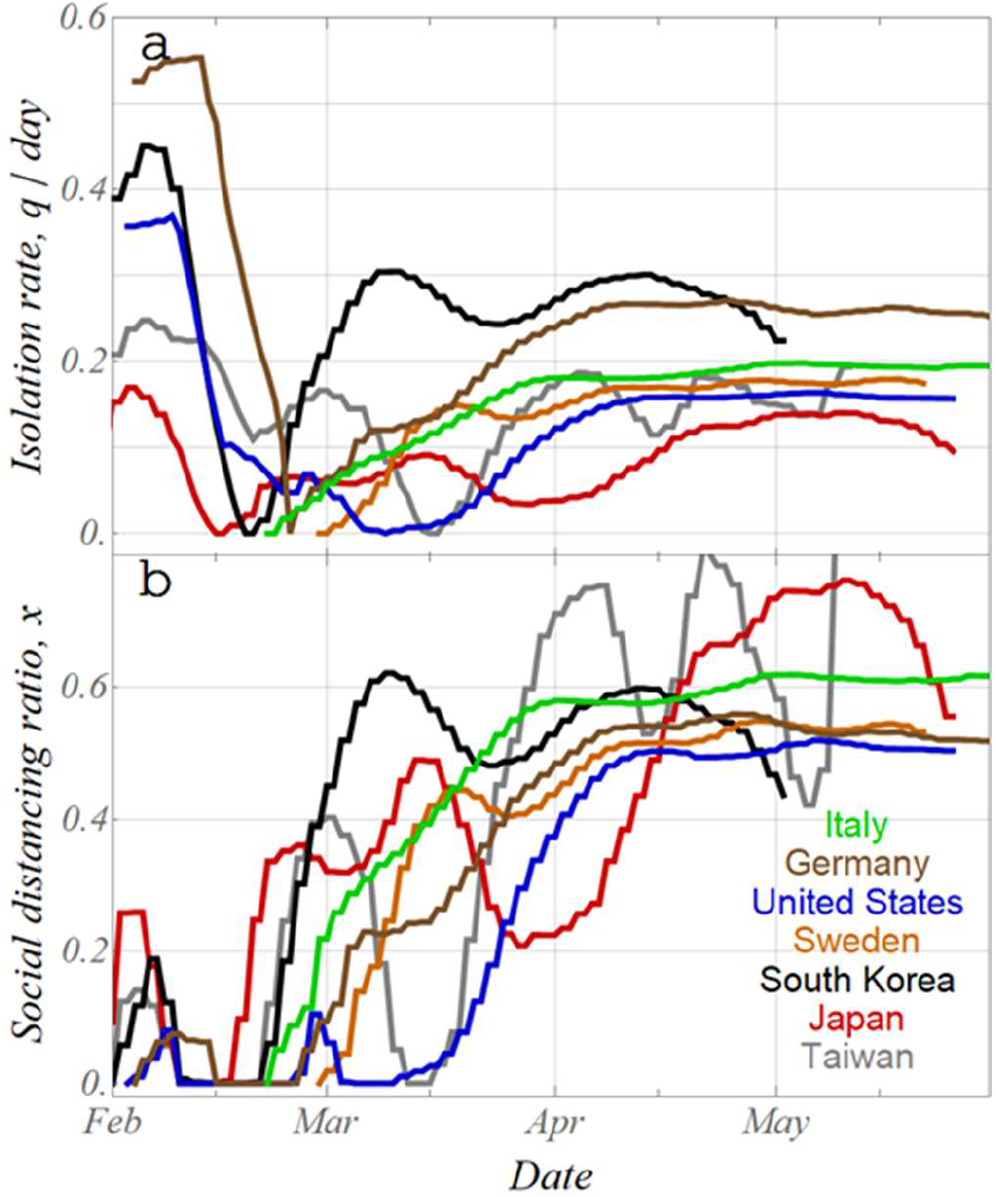
Daily variation of isolation rate q and social distancing ratio x for 7 countries

The variation in both of q and x for Japan are described in Figure 4 as an example. For the sake of explanation, daily λ value curve in Fig. 1 and the corresponding equi-λ lines calculated by eq. (5) are shown in Fig. 4.

1. Starting from q=0.1 at which the horizontal axis (x=0) intersects with the equi-λ line (λ=0.1, Fig. 4b) corresponding to λ=0.1 at the start of infection (Fig. 4a).
2. Then, the daily λ suddenly decreases and becomes 0 (Fig. 4a), both q and x increase slightly to the position intersecting the equi-λ line (λ=0).
3. Thereafter, when the daily λ increases and reaches λ_max_ (Fig. 4a), the q-x locus reaches the origin (∆λ = 0).
4. After reaching λ_max_, the locus on q-x moves to the upper right as daily λ decreases with fluctuation.
5. At the point when daily λ cuts the line of 0 and turns negative (mid-April in Fig. 4a), it moves to the point where it intersects with the equi-λ line (λ = 0) again (this point corresponds to the maximum number of new cases).
6. During the convergence stage (daily λ<0), q-x does not change significantly and reaches the final convergence point where it intersects with the equi-λ line (λ=-0.05) or less.

**Fig. 4.**
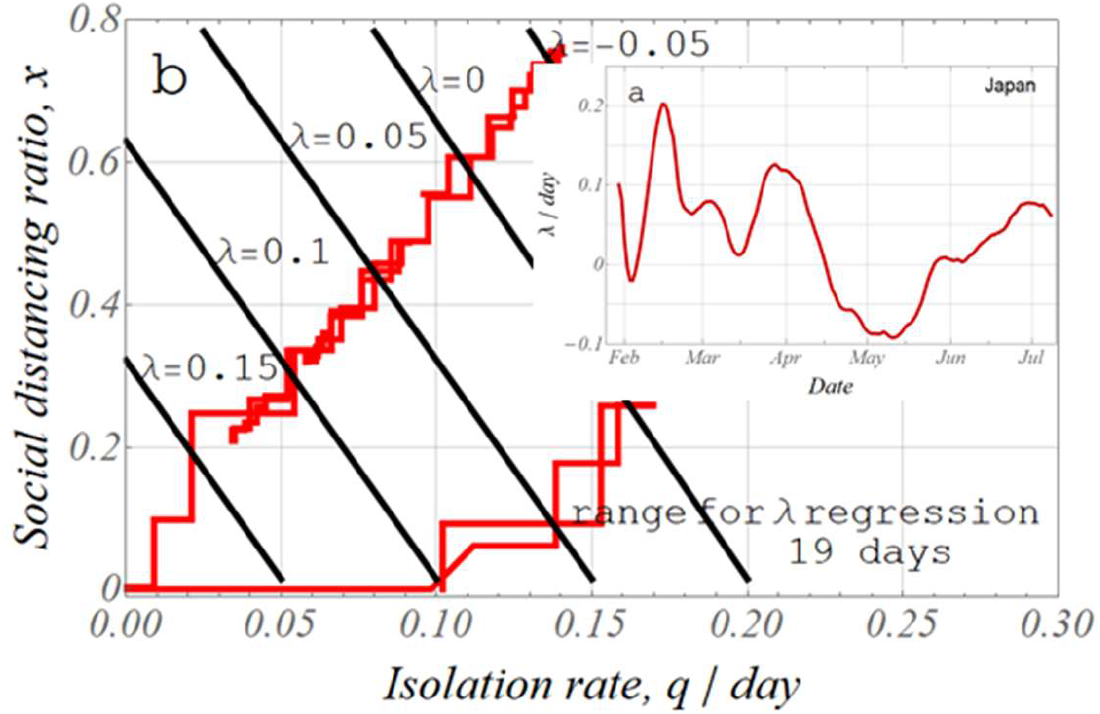
Moving locus of (q,x) point on the q-x plane, explaining the method of bipedal walk sequential calculations referring Fig.4a in case of Japan

In this way, the isolation rate q and the social distancing ratio x follow a complicated locus on the q-x plane corresponding to the variation in the daily λ value after the start of infection. Figure 5 shows the q-x loci of 7 countries. In the figure, the day points for the peak are indicated by dots.

**Fig. 5.**
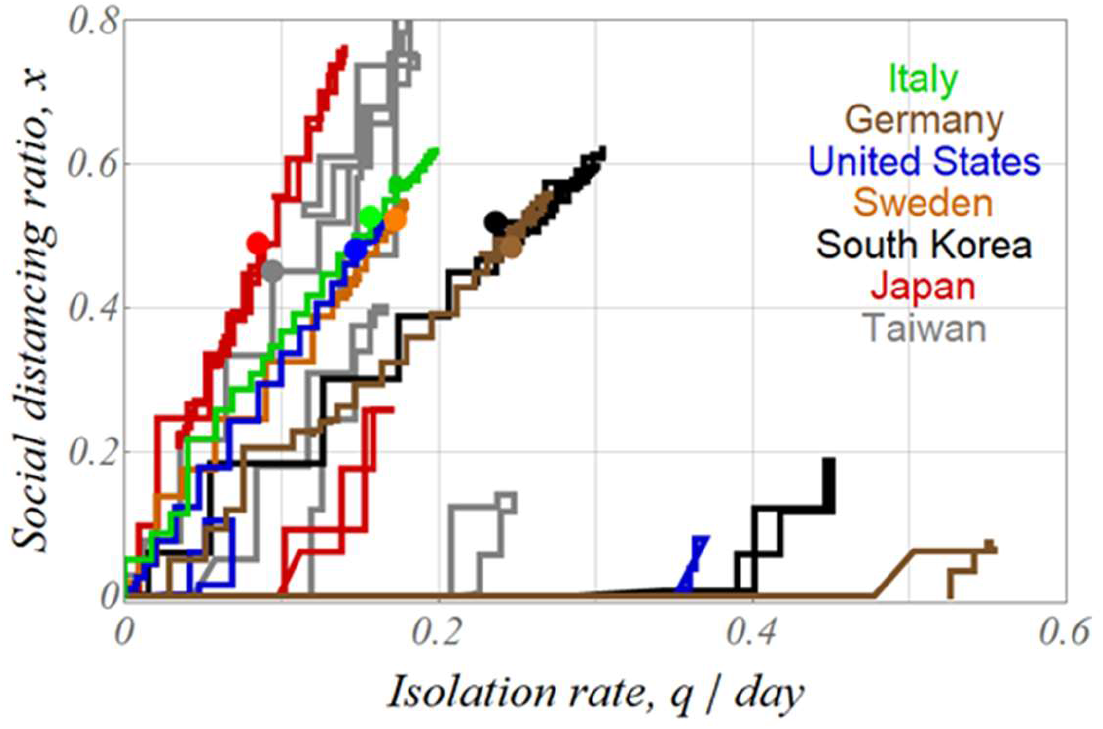
Moving locus of (q,x) point on q-x plane for 7 countries; dots marked at peak day

Keeping in mind the common locus starting from on the q-axis as revealed in Fig. 4, Fig. 5 clearly shows the differences in q-x relationships between countries. In contrast to Japan and Taiwan, where the variation in isolation rates is small, the magnitude of the variation in isolation rates in Germany, Korea and the United States is noteworthy. In the cases of Sweden and Italy, λ_max_ was reached on the first day of infection (Fig. 2b), so their locus started from origin on the q-x plane. There is also a large difference in the straight line up to the right during the spread of infection to the convergence period. In Japan and Taiwan, the isolation rate remains low and the social distancing ratio increases, meanwhile in Germany and Korea the isolation rate increases significantly and the social distancing ratio also increases. The United States, Sweden and Italy fall somewhere in between.

From these behaviors, the actual infection process can be interpreted as follows. At a very beginning of infection, when the number of infected persons is small, the isolation rate is high because fewer tests are needed to efficiently find and isolate infected persons. Then after, the further increase in the number of infected persons makes test and isolation unable to catch up with and the balance between the increase in the number of infected persons and isolation is lost, the number of infected persons rapidly increases to λ_max_ point and both q and x decrease. After that, at the stage of catching up by tests with the increase in the number of infected persons (until the maximum number of infected persons is reached), the effects of both countermeasures of isolation and social distancing are maximized, thereafter the number of infected persons decreases steadily.

Since the calculated isolation rate and social distancing ratio are strongly influenced by the degree of variation in daily λ values, care should be taken in setting the regression range of daily λ values. Taking Japan as an example, Figure 6 shows the relationship between daily λ value and the isolation rate and social distancing ratio when the regression range of daily λ values is changed from 7 to 19 and 31 days. As the regression range expands, the fluctuation in daily λ value becomes smaller, and the position of λ_max_ shifts almost to the first day of infection (Fig. 6a). The proximity of λ_max_ day to the first day of infection means that the q-x locus curve starts near the origin, and this behavior is similar to those of Sweden and Italy, as described above.

**Fig. 6a,b.**
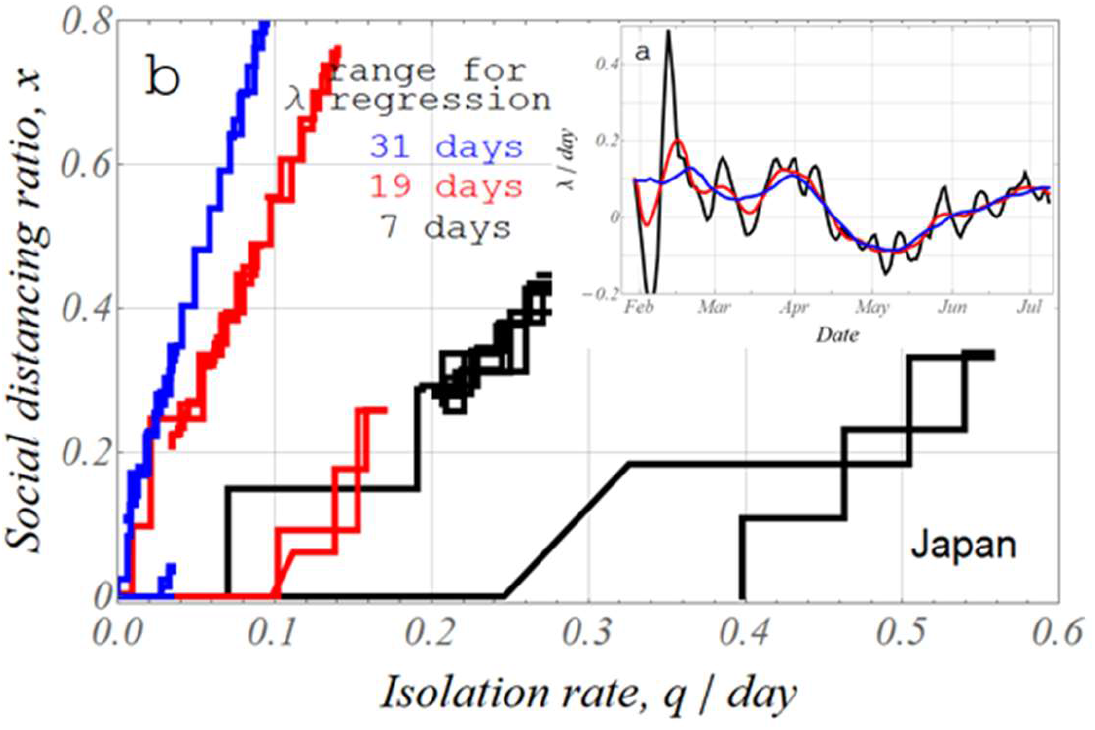
Effect of the range of days for regression calculation of daily λ on (q,x) locus for Japan

## Importance of the specific value βN for each country

The above analysis reveals that the magnitude of λ_max_ affects the varying degree in q and x through the magnitude of the variation in the left-hand side of eq. (5). This suggests that the effect of the transmission coefficient βN (=λ_max_ + γ), which is specific value for each country, on the variable width of q and x, and thus on the degree of influence of the countermeasure differs. In Japan and Taiwan, where λ_max_ (βN) is small, the spread of infection is slow, while in other countries where λ_max_ (βN) is large, the spread of infection is rapid.

In order to examine the effect of βN on the spread of infection, the relationship between λ_max_ and total cases for the other many countries was examined. The latter were determined at the convergence day, which is more than 1 week before the last day of the database. The relationship of both is shown in Figure 7a. The calculation of both parameters were performed for about 100 countries with more than 400 new cases or more than 1 million populations among all 211 countries/regions in the database. Although there is a large variation, the lower λ_max_, the lower the upper limit of the total cases. This means that even at the same λ_max_ the total cases fluctuate widely, and the range of fluctuation increases with λ_max_. In fact, data for the seven countries were denoted in red in Fig. 7, the total cases in five countries with λ_max_ > 0.3 increases from the smallest in South Korea to the smaller in Germany, then Italy, Sweden, and the U.S., in the order in which the total cases increase according to the so-called national measures. The countries with lower total cases at fixed λ_max_, are considered to have taken effective measures within the range of λ_max_, that is, q and x, which are allowed by the specific value βN (λ_max_), and as a result, the total cases is considered to have been controlled at a low level.

**Fig. 7a,b.**
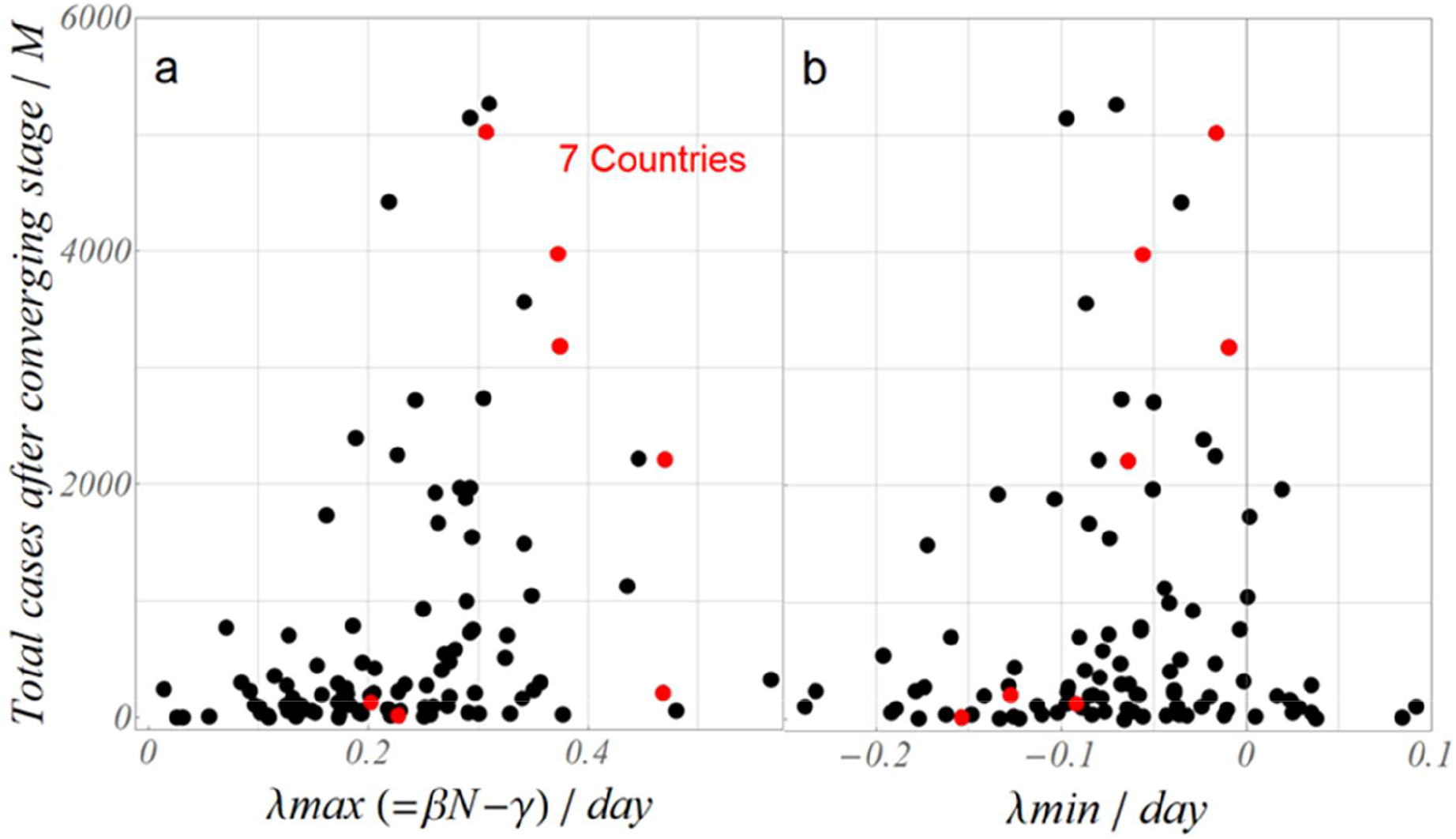
Relationship between total cases after converging stage and *λ_max_* (=*βN* − *γ*) *λ_min_* for more than 100 countries; Shown in red dots are Japan, Taiwan at around *λ_max_* 0.2, and for *λ_max_* > 0.3, South Korea, Germany, Italy, Sweden and USA in the order of increasing total cases.

In Fig. 7b, which is organized by λ_min_ instead of λ_max_, the lower λ_min_ (the larger the negative value), the lower the upper limit of the total cases. In other words, lower λ_max_ and lower λ_min_ are effective in reducing infection, which corresponds to the self-evident fact that the increasing rate of infection must be lower (low λ_max_) and the decreasing rate must be higher (low λ_min_).

Through the above examination on the relationship between the transmission coefficient βN and the total cases, the importance of the former was confirmed. As described in previous section, the transmission coefficient βN was closely related with λ_max_ as a specific value for each country, which was assumed to be constant throughout the infection period. Although detailed examination of this coefficients is beyond the scope of this study, it can actually be interpreted as specific values that represent the unique characteristics of each country and society, including such as the lifestyle of interpersonal activities, and further research is needed.

## Conclusions

For the purpose of verifying the validity of the newly proposed SIQR model, in which the exponent λ of the exponential function expressing the number of newly tested positive persons was defined as an linear equation explicitly with three important parameters, transmission coefficient, social distancing ratio x and isolation rate q, the trend of the number of isolated persons in publicly available database for COVID-19 epidemic were analyzed. These analyses were performed for 7 countries including Japan, Taiwan, South Korea and western countries.

Results are summarized as follows.

- Daily λ values were regression analyzed, and were shown to vary from positive value during the increasing stage of infection, then decrease through 0 (peak stage) to negative value (convergence stage). During this epidemic period, λ reaches maximum value λ_max_ which is closely related to the transmission coefficient βN.
- The isolation rate, q and the social distancing ratio, x were calculated by sequential calculations using the equation relating daily λ with the isolation rate and the social distancing ratio, and the locus of q-x relation with the progress of infection were determined.
- Japan and Taiwan exhibit higher social distancing ratio, South Korea and Germany show higher isolation rates, and Italy, Sweden, and the United States show intermediate characteristics.
- Transmission coefficient βN is lower in Japan and Taiwan, higher in 5 other countries. The cumulative number of isolated persons after convergence tends to decrease as the transmission coefficient becomes lower, which were confirmed in examination for more than 100 countries.

## Data Availability

Only publicly available database was used.

https://ourworldindata.org/coronavirus-source-data

## Acknowledgment

The author would like to thank professor emeritus Takashi Odagaki at Kyushu University for several comments and especially his SIQR model that inspired the author to start this analysis, and also professor emeritus Ikuo Yoshihara at University of Miyazaki for detailed and fruitful comments and discussions in preparing this paper.

## References

1) W.O. Kermack and A.G. McKendrick, Proc. Ry. Soc. A115(1927) 700

2) J. Makino, Science (Iwanami Shoten) 90(2020) 428, in Japanese publication

3) T. Odagaki, Bussei Kenkyu@Web Vol.8, No.2.082101(2020), in Japanese, T. Odagaki, https://doi.org/10.1101/2020.06.02.20117341

4) Our World in Data COVID-19 dataset, https://ourworldindata.org/coronavirus-source-data, (Accessed: 2020.7.12)

